## Supplementary material for "Enabling large-scale screening of Barrett’s esophagus using weakly supervised deep learning in histopathology"

### 1 Study overview

**Table 1** Key study elements of our paper listed according to recent reporting guidelines by[1] and[2] for applications of machine learning (ML) in clinical research.

| | |
|---|---|
| **Study design** | |
| Clinical question | Can we detect Barrett's esophagus (BE) from the routinely-stained hematoxylin and eosin (H&E) slides using weakly-supervised deep learning methods? |
| Model task and outputs | Binary classification of H&E whole-slide images into BE positive or negative. Slide attention heatmaps showing high and low attended tiles by the model to make the predictions. |
| Intended use of results/target user | Predictions and model outputs (e.g., slide attention heatmaps) can be made available to clinical supporters as part of the Cytosponge-TFF3 test management, for instance, to assist pathologists in semi-automated workflows (see Results), enhance the scalability of the test, and improve patient outcomes. |
| **Study population and setting** | |
| Population | Comprises of data from patients attending Barrett's surveillance or screening programs as enrolled in one of the two studies, namely, DELTA and BEST2 (see Methods: Discovery and evaluation external datasets). |
| Study setting | DELTA implementation study and BEST2 clinical trials registered in the UK (see Methods: Discovery and evaluation external datasets). |
| Data source | Cyted Ltd, Cambridge, UK. |
| Cohort selection (exclusion and inclusion criteria) | All available slides from the DELTA study. Licensed slides available to Cyted Ltd from the BEST2 study. |
| **DELTA Patient demographics** | |
| Age (Median, IQR | 66 years (IQR 58-74). |
| Sex | M (n=726): F (n=508): Missing (n=25). |
| Ethnicity | Not provided. |
| Socioeconomic status | Not provided. |
| **Data sources** | |
| Data types | Whole-slide images in NDPI format (discovery dataset) and SVS format (external dataset), converted into TIFF at objective magnification 10× (0.92 $\mu$m/pixel). Tiles generated of size 224 × 224 pixels ($\approx$ 200 × 200 $\mu m$) from whole-slide images of sizes of order of $1 \times 10^8$ pixels. Structured metadata in TSV format. |
| Data collection and processing | See Methods: Discovery and external evaluation datasets, Data preprocessing. |
| Data structures | RGB image (array of 3 channels), binary label for BE. |
| Data partitions | See Fig. 1c |
| **Model architecture and training** | |

| | |
|---|---|
| ML method and rationale | Weakly supervised multiple instance learning (MIL) method can achieve a high diagnostic performance to detect BE from H&E slides due to a strong alignment with the nature of the task: a slide is labeled BE positive when goblet cells are detected in a small area of the whole-slide image — this is a classical MIL task and resembles the assessment criteria of expert histopathologists. We use a weakly supervised deep learning network architecture inspired by Transformer-MIL proposed in[3]. The resulting model architecture is called BE-TransMIL. Benchmarked encoders: ResNet18, ResNet50, DenseNet121, Swin-T (see Methods: Model architecture). |
| Features | Learnable features selected by the deep learning model. Interpretability analysis highlights the following features with higher attention values given by the model (see Results, Fig. 2, Fig. 3, Fig. 5).<br>H&E slides: Mucin-containing goblet cells are visible with a distinct cellular morphology in the high-attention tiles.<br>trefoil factor 3 (TFF3) slides: Goblet cells show positive staining of the brown histochemical stain in the high-attention tiles. |
| Data labels (Gold standard) | The BE positive or negative labels are derived from pathologists' diagnostic reports. Pathologists visually assessed each slide pair (H&E, TFF3) to make their BE diagnosis. |
| Missingness | Not applicable |
| Hardware, software, packages | • Azure ML infrastructure (https://azure.microsoft.com/) for pre-processing TFF3 slides, training and evaluating deep learning models<br>• HistoQC configv2.1[4] for pre-processing H&E slides<br>• MONAI[5] for data pre-processing and tiling-on-the-fly<br>• 8 NVIDIA V100 GPUs for training, 1 NVIDIA V100 GPU for inference<br>• 40 CPU cores for tiling on-the-fly<br>• Python 3.9 for code implementation<br>• PyTorch 1.1.0[6] and PyTorch-Lightening 1.6.5[7] for implementing the deep learning models and model evaluation<br>• Scikit-learn[8], Scikit-image[9] for data processing and analysis<br>• SimpleITK[10] for registration of H&E and TFF3 slides |
| Data split | 80:20 split of discovery dataset into development (training, validation) and test datasets (See Fig 1c). Four-fold cross validation leading to 60:20:20 split over the entire discovery dataset. Validation and test datasets were randomly selected, stratified according to distributions of class labels and patient pathway (surveillance or screening). |
| Model parameters and hyperparameters | BE-TransMIL model architecture: ResNet50 encoder + 4 Transformer encoder layers + attention MIL pooling + multi-layer perceptron (MLP) classifier.<br>Hyperparameter tuning of models based on classification accuracy, prioritized on specificity (in order to identify negative cases with high confidence for screening populations). |

| | |
|---|---|
| Model training | BE-TransMIL models trained for binary classification task using:<br>• BCE loss, ADAM optimizer ($\beta_1 = 0.9, \beta_2 = 0.99$)<br>• Batch size = 8<br>• Bag size = varying (ResNet18: 2300, ResNet50: 1200, Swin-T: 1100, DenseNet121: 700),<br>• Hidden dimension of attention pooling layer = 2048<br>• Number of epochs = 50<br>• Learning rate = 3e-5<br>• Weight decay = 0.1<br>• Dropout=None<br>• Random seed = 42<br>• Scalability methods: Activation checkpointing (training), encoding in chunks (whole slide inference). See Methods: Model description for details. |
| **Model evaluation/validation** | |
| Evaluation measures (metrics) | AUROC, AUPR, Accuracy, Sensitivity, Specificity (at threshold=0.5) for comparing and model selection, with priority to AUROC and AUPR as these are threshold-agnostic metrics.<br>Accuracy, AUROC, AUPR, Sensitivity, Specificity at selected operating point (corresponding to 0.85 validation sensitivity for clinical utility) for reporting performance on discovery and external test sets.<br>ROC curves with bootstrapping for 95% confidence intervals (CI). |
| Internal model validation | Discovery dataset comprises of cases from the DELTA implementation study.<br>For each fold in four-fold cross-validation, discovery training set consists of 912 slides and validation set of 229 slides for model selection and comparison.<br>Discovery test set consists of 229 slides for model evaluation. |
| External model validation | External dataset comprises of slide images from the BEST2 case-control clinical trial.<br>The external validation set consists of 725 slides. |
| Interpretability analysis | We analyze the attentions of the model qualitatively and quantitatively, correlate the findings with TFF3 stain for which goblet cells show positive staining, and analyze failure modes to ensure model outputs are interpretable (See Results). The analysis includes:<br>• Visual assessment of slide attention heatmaps and top/bottom attention tiles to analyze the visual features selected by the models to make a decision.<br>• GradCAM saliency maps to analyze fine-grained attention in tiles.<br>• Stain-attention correspondence analysis to correlate the attentions with TFF3 stain ratio.<br>• Failure modes analysis to assess the failures of shared and individual mistakes of H&E and TFF3 models. |

| | |
|---|---|
| Transparency, reproducibility, code re-use | Data cannot be shared by corresponding author due to license agreements of Cyted Ltd with partners. The study protocols for DELTA and BEST2 are publicly available. Data may be available upon request to the original institution. All data used was deidentified. The open-source repository https://github.com/microsoft/hi-ml/tree/main/hi-ml-cpath contains the code and library requirements for data preprocessing, core BE-TransMIL network architectures for training and evaluation of deep learning models, and statistical analysis presented in this study. More detailed code will be added to this repository before publication. |
